# Assessing Cerebral Visual Impairment Through Non-Verbal Responses: Protocol Development and Feasibility of EchoCue™ [Registered Report, stage 1]

**DOI:** 10.1101/2025.06.18.25329869

**Authors:** Edwige Smague

## Abstract

This Stage 1 Registered Report outlines the protocol for a feasibility study of EchoCue™, a novel paradigm designed to assess cerebral visual impairment in children aged 2 to 6, through non-verbal response patterns. The paradigm minimises the reliance on verbal instruction or active cooperation, making it suitable for populations traditionally considered difficult to assess. The method pairs auditory and visual stimuli with the recording of reflexive responses, such as gaze shifts, blinks, and startle reactions, using an integrated infrared eye-tracking system. The primary objective of this study is to assess the feasibility of the data collection procedures and to generate robust estimates of effect sizes with 95% confidence intervals for the protocol’s core measures, in order to inform the design of a future large-scale confirmatory trial.

## Introduction

Cerebral visual impairment (CVI) is a major cause of childhood disability in Australia and high-income countries. Most existing screening tools rely on task-based protocols requiring active participation, rendering them inaccessible to the most severely affected children (1). The difficulty is compounded by the neurological origin of CVI involving damage to the visual pathways or the cortex, therefore necessitating evaluation methods that differ from traditional ophthalmic assessments (2).

This clinical challenge is further amplified by the symptomatic overlap with other neurodevelopmental disorders, such as autism spectrum disorder (ASD), including peculiarities in eye contact, stereotypical behaviours, and atypical responses to sensory stimuli, which can delay or even prevent the correct identification of visual impairments, thus depriving these children of appropriate and early care (3). There is, consequently, an urgent clinical and scientific need for an evaluation methodology capable of objectively testing the foundational sensory processes that underpin a child’s perceptual abilities (4). Such an approach must minimise reliance on the child’s active participation to provide objective markers of neurosensory integrity. EchoCue^TM^ has been designed as an objective screening and assessment tool intended to provide critical evidence within a comprehensive clinical evaluation.

Developed in this context, EchoCue^TM^ represents a new methodological approach based on the presentation of calibrated sensory stimuli (auditory and visual) and the synchronised capture of reflex responses (eye movements, blinks, startles) using an integrated eye-tracking system and a motion sensor. While the primary aim of this study is to assess the method’s initial feasibility, future studies with larger cohorts will be required to establish normative data.

## Method

EchoCue™ is an original screening framework designed by Edwige Smague in 2025 as part of a doctoral research initiative. The device is not yet commercialised and is currently under academic development for feasibility testing in Australia. The methodology presented in this article is based on the EchoCue™ conceptual framework, which enables the objective screening of reflexive and bottom-up neurovisual functions using strategically calibrated multimodal sensory stimuli.

### Sample Size Justification

The planned sample size (n=15 per group) is determined by recruitment feasibility for this initial proof-of-concept study. A brief internal feasibility check (n=5 per group; CVI, ASD, TD) will be conducted at the outset to confirm task tolerability, optimise calibration, and verify data-collection logistics before full recruitment. Although this study is exploratory, it is designed to detect a large effect size (Cohen’s d ≥ 0.8), as a clinically useful screening tool must be sensitive to clear, robust differences in neurosensory function. Data from this study will provide effect size estimates to inform power calculations for future validation trials designed to meet stricter statistical thresholds (e.g., 90% power). The primary goal of this study is to establish the feasibility of the protocol and generate robust estimates of effect size and confidence intervals (CIs) to guide the design of a future confirmatory trial. If the preliminary feasibility check indicates poor tolerability, calibration instability, or insufficient data capture quality, protocol parameters (e.g., stimulus intensity, trial duration, eye-tracking calibration procedures) will be modified accordingly, and feasibility will be re-verified before progressing to full recruitment. A preliminary power simulation (SIMR) indicates an estimated power of approximately 0.70 for the planned comparison, consistent with the exploratory aim of this Stage I design.

### Power Simulation

A power simulation using the SIMR package (Green & MacLeod, 2016) indicated an estimated statistical power of approximately 0.70 for the planned LMM analyses. As this value falls below conventional thresholds for confirmatory inference, the present study is therefore considered exploratory and estimation-focused.

### Participants

A total of 45 participants, within the preschool age range of 2 to 6 years, will be recruited to ensure developmental homogeneity for this initial feasibility study. Participants will be divided into three age and sex-matched groups (n=15 per group): (1) children diagnosed with CVI; (2) a clinical control group with a diagnosis of ASD and no known visual impairment; and (3) a typically developing control group. All participants’ adaptive functioning will be assessed and statistically controlled for (see the Measures and Statistical Model specification sections).

### Feasibility Metrics

In line with the primary goal, feasibility will be evaluated using three primary indicators:

1. **Completion Rate**: The proportion of participants who complete all core testing blocks (1–3).
2. **Tolerance**: Observational coding of behavioural signs of distress (e.g., crying, agitation) during testing.
3. **Parents’ Acceptability**: A brief post-session questionnaire rating the perceived comfort and ease of participation on a 5-point Likert scale (1=very low, 5=very high).

Items assess:

‒ Perceived comfort of the child during the task
‒ Clarity of instructions provided to caregivers
‒ Ease of participation
‒ Acceptability of session duration
‒ Willingness to participate again or recommend the procedure.

The mean score will be calculated per participant, with high acceptability defined as ≥4/5 at the group level.

Feasibility will be supported when ≥80% of participants complete all blocks, <10% discontinue due to distress, and the mean parent acceptability score ≥4/5.

### Recruitment and Inclusion/Exclusion Criteria

Participants will be recruited from key paediatric institutions and support networks in Brisbane, Australia, including the Queensland Children’s Hospital (QCH), Mater Children’s Private Brisbane, National Disability Insurance Scheme (NDIS), Vision Australia, Autism Cooperative Research Centre (Autism CRC), Next Sense, Statewide Vision Impairment Services (via the Department of Education Queensland), Queensland University of Technology (QUT), and The University of Queensland (UQ).

This broad recruitment network, tapping into major clinical and community support services, provides a realistic basis for achieving the recruitment target of 15 per group for this specific population. This target appears highly feasible; for context, a recent study at just one of these Australian tertiary referral centres successfully recruited 102 children with CVI or related neurodevelopmental delays for a cross-sectional study (5).

Inclusion criteria for the CVI group will include: (1) a formal clinical diagnosis of CVI from a qualified professional (e.g., paediatric neurologist or ophthalmologist), confirming the visual impairment is of neurological origin; and (2) an age between 2 and 6 years.

Inclusion criteria for the ASD group will include: (1) a formal clinical diagnosis of autism spectrum disorder from a qualified professional (e.g., paediatrician, psychologist), based on established criteria such as the DSM-5; (2) no known or suspected visual impairment of either ocular or cerebral origin; and (3) an age range between 2 and 6 years.

Inclusion criteria for the TD group will include: (1) no known developmental, neurological, or psychiatric diagnoses; (2) no parental or clinical concerns regarding vision, hearing, or general development; (3) born full-term with no significant perinatal complications; and (4) an age range between 2 and 6 years.

Notably, there will be no exclusion criterion based on cognitive or intellectual level (e.g., IQ score <70). A key objective of this study is to develop and assess the feasibility of a tool for the full range of children with CVI, including those with co-occurring intellectual disability.

Exclusion criteria for all groups will include: (1) the presence of a severe, uncorrected ocular pathology that could primarily account for visual deficits; (2) a diagnosed severe or profound hearing impairment that would prevent perception of the auditory stimuli; or (3) an unstable medical condition precluding participation in a 15-minute session.

Written informed consent will be obtained from the parents or legal guardians of all participants.

## Measures

### Developmental and Adaptive Functioning

To assess the developmental and adaptive functioning level of each participant, parents or legal guardians will complete the Vineland Adaptive Behavior Scales, Third Edition, Vineland-3 (6).

The Vineland-3 is a standardised assessment completed by parents or primary caregivers that evaluates communication, daily living skills, socialisation, and motor skills. The Adaptive Behavior Composite score will be used as a covariate to control for developmental differences across groups.

This measure was deliberately selected because, as a parent-report tool, it minimises the performance-based demands that are often prohibitive for young or clinically affected children, thereby aligning with the protocol’s feasibility objective. Although the Vineland provides an indirect rather than performance-based index, it represents a robust and pragmatic method for statistically controlling overall developmental level in this population.

### Theoretical Rationale for Using Auditory Probes to Assess Neurovisual Function

The EchoCue^TM^ protocol is based on an objective assessment method that tests the child’s neurosensory functions without the need for the child to understand instructions or actively participate (7). The approach involves exposing the child to a series of finely calibrated auditory and visual stimuli while synchronously recording their involuntary reflex responses (8), such as blinks, micro-saccades, or startles, and visual orientation responses.

A key element of this method is using a wide range of sound frequencies, from 20 to 8000 Hz. Each frequency range allows distinct neurological circuits, both subcortical and cortical, to be explored in order to better understand how the brain perceives, integrates, and responds to multisensory stimuli (9). Low-frequency sounds (20—250 Hz) act as a subcortical marker. This range is particularly relevant for assessing reflexive response in non-instructional paradigms (10).

These frequencies are perceived by the human body as both vibrotactile stimulation and sound. They are transmitted partly by bone conduction, directly stimulating the cochlea and vestibular system, thus bypassing any disorders of the outer or middle ear (11). More important still, they are processed primarily by the archaic neural circuits of the brainstem due to their simple acoustic configuration and their strong ability to arouse sensory alert systems (12, 13). They are particularly effective at triggering involuntary reflexes managed by the brainstem, such as the stapedius reflex (an involuntary auditory reflex that protects the inner ear from loud sounds) and the startle reflex (14, 15). The use of these frequencies thus makes it possible to measure a basic neurological response, a sign of the integrity of the subcortical pathways, even before the information reaches the higher cortical areas. This approach is consistent with findings from auditory neuroscience showing that low-frequency auditory stimuli can promote autonomic regulation, including parasympathetic activation, through mechanisms such as vagal nerve modulation (16). These effects have been associated with reduced anxiety and increased physiological markers of calm in both clinical and developmental populations (17, 18).

Beyond their function as a subcortical probe, the use of these low-frequency sounds is also informed by emerging research on their potential therapeutic effects (18). Studies have suggested that certain frequencies can have a regulating or calming effect on the nervous system (19), which may help in managing anxiety in paediatric populations, including children with ASD during stressful procedures (20). This dual function, as both an objective probe and a potentially regulating stimulus, is a key feature of the EchoCue™ protocol’s design to maximise participant comfort.

Medium to high frequencies (250—8000 Hz) are used for cortical analysis. This range is essential for assessing the classical auditory pathway and higher cognitive functions (21, 22). It is necessary for multisensory integration tasks, where the brain has to compare and merge auditory information with visual information (23). Difficulty processing these sounds or integrating them correctly with a visual stimulus may reveal dysfunction in associative cortical areas (e.g., the temporoparietal junction), which are often involved in CVI (24).

To establish a reliable baseline of involuntary orienting behaviour, the EchoCue™ protocol begins with an auditory-only condition (Block 1) using lateralised, high-intensity sounds (25). This condition is not intended to elicit voluntary saccadic eye movements alone, but rather to evoke a multimodal orienting reflex that includes head turning (26), postural adjustment, and, in some cases, eye movement (27). This reflex is mediated by the superior colliculus (28), a subcortical structure that integrates spatial auditory and visual inputs and directs attention and movement toward salient stimuli (29), even in complete darkness (30). The orienting response to sound occurs early in development and does not rely on cortical-level processing or explicit behavioural intention (31). The measurement system is specifically designed to capture this multimodal response. The TrueDepth facial tracking system (iPad Pro) captures subtle gaze and head movements, while a millimetre-wave (mmWave) radar device tracks full-body posture and micro-adjustments in real time. Together, these technologies provide a robust, non-invasive index of the brain’s responsiveness to lateralised sound.

The extent to which a simultaneous visual stimulus (Block 3: Congruent Audiovisual condition) modulates this auditory-driven orienting response serves as an indirect index of functional visual processing. This approach captures the visual modulation of a non-visual reflex, offering an objective, instruction-free marker of visual function, particularly valuable in CVI, where conventional behavioural measures are often unavailable.

### The Standardised Screening Protocol

The test session is brief, lasting between 12 and 15 minutes. It is structured in sequential blocks (see Table 2). While the broader EchoCue^TM^ framework is designed to assess responses across a 20—8000 Hz range, this initial study uses a key subset of these frequencies (500 Hz and 2000 Hz) to address the pre-registered research questions.

### Safety and Distress Protocol

Participant safety and comfort are paramount. All auditory stimuli will be calibrated and will not exceed a maximum intensity of 70 dB SPL, a level considered safe for brief exposures in paediatric populations (32).

To ensure auditory safety in the event of positional variation, participant distance from the device will be visually monitored throughout testing. Although stimuli are calibrated to 70 dB SPL at the head position, even substantial forward movement is expected to increase intensity only moderately (approximately +6dB per halving of distance), remaining within accepted short-duration paediatric safety limits. Sessions may be paused and seating position adjusted if a child approaches the speaker closely enough to materially alter sound exposure.

Throughout the session, the researcher will continuously monitor the child for any signs of distress (e.g., crying, agitation, prolonged gaze aversion). The protocol includes flexible breaks between blocks. The session will be terminated immediately if the child exhibits clear signs of distress that cannot be readily soothed, or if the parent/guardian requests discontinuation.

### System Architecture and Measurement Parameters

The implementation of this protocol requires a simple, accessible, and non-invasive technological platform capable of fulfilling three essential functions: stimulus generation, eye-tracking, and motion detection.

### Stimulus Generation and Primary Device

A central unit, an iPad Pro (model 2021 or later), generates the protocol’s audio-visual stimuli. Stimulus presentation and data acquisition are managed by a custom-built application developed in Swift using Apple’s ARKit API (version 5 or later)(33).

### Eye-Tracking and Saccadic Reaction Time (SRT) Measurement

To ensure reliable data capture, the protocol uses the iPad’s integrated infrared TrueDepth camera system. This system operates at a 60 Hz frame rate, providing a temporal resolution of 16.7 ms, which is considered sufficient for reliably measuring saccadic reaction times. Furthermore, it is coupled with the ARKit API, which offers the significant advantage of simultaneous head and face tracking, allowing for the automatic correction of gaze for head movements.

Gaze data is used to calculate Saccadic Reaction Time (SRT), defined as the time elapsed between the onset of a visual target and the onset of the saccade towards it (34).

◾ **Saccade Definition**: A saccade is detected using a velocity-based algorithm. An eye movement is classified as a saccade when its angular velocity exceeds a threshold of 30° per second and is considered complete when the velocity drops below this threshold. This distinguishes saccades from slower movements like ocular drift.
◾ **Onset Definition**: The onset of the saccade is the first video frame at which the eye’s angular velocity exceeds the 30°/s threshold.
◾ **Valid Timeframe**: For an SRT calculation, a saccade’s onset must occur between 150 ms and 700 ms after the visual target appears.

### Motion Detection and Startle Amplitude Measurement

A single, external mmWave radar sensor (e.g., Acconeer XM122) is used to quantify fine motor responses from a distance of at least 60 cm (35). Given the novelty of this technique in a paediatric clinical context, this measure is considered exploratory.

The radar signal is used to measure the Startle Amplitude, which is operationally defined as a rapid, involuntary flinch in response to an auditory stimulus.

◾ **Measurement**: The radar provides a continuous measure of motion energy. The Startle Amplitude for a given trial is the peak motion energy recorded within a pre-specified time window.
◾ **Timeframe**: The analysis window for detecting a startle is from 40 ms to 200 ms following stimulus onset.
◾ **Units and Normalisation**: The value is in arbitrary units (a.u.) of motion energy. It is normalised by dividing it by the standard deviation (SD) of the motion energy recorded during a 100 ms silent baseline period immediately preceding the stimulus.
◾ **Threshold**: A response is classified as a valid startle reflex only if its peak amplitude exceeds three SD above the mean of the pre-stimulus baseline energy. This startle-detection criterion is conceptually distinct from the trial-level exclusion rule, which uses the same 3 SD threshold solely to identify excessive pre-stimulus head movement.

### Stimulus Presentation Protocol

The pre-registered research questions are addressed using data from Blocks 1, 2, and 3 (see Table 2). The details for these blocks are as follows:

◾ **General Parameters**: Auditory stimuli will be presented via the iPad’s internal stereo speakers, calibrated to deliver a peak intensity of 70 dB SPL at the participant’s head position. Visual stimuli are a white LED flash (40 ms duration) presented on the left or right side of the screen. Each trial begins with a 500 ms central fixation point, and the inter-trial interval will vary randomly between 1500 ms and 2500 ms.
◾ **Block 1 (Auditory-only)**: Consists of 20 trials (10 left, 10 right). For the trial relevant to the core research questions, a subset of the available frequencies (500 Hz and 2000 Hz) will be presented in a randomised order.
◾ **Block 2 (Visual-only)**: Consists of 20 trials (10 left, 10 right), presented in a randomised order.
◾ **Block 3 (Congruent Audiovisual)**: Consists of 20 trials (10 left, 10 right). The auditory frequency (500 Hz or 2000 Hz) and location will be randomised. The visual flash will always be presented at the same location as the sound.

### Data Quantification and Analysis

In line with the system’s philosophy, the EchoCue^TM^ software does not provide an interpretation or diagnosis. Its role is to act as a quantification engine that applies signal processing algorithms to transform raw data from the various sensors into the objective metrics required for analysis, as detailed in the System Architecture and Measurement Parameters section. These quantified data are then compiled into a clinical report, allowing the practitioner to base their evaluation on objective and reproducible evidence (36).

The pre-registered research questions outlined in Table 1 are directly linked with specific blocks of the EchoCue^TM^ protocol. Data from Blocks 2 and 3 will be used to estimate group differences in Saccadic Reaction Time (SRT), while data from Blocks 1 and 3 will be used to estimate group × condition effects on Startle Amplitude. Data from Blocks 4 and 5 are collected for documentation and potential future exploratory validation but are not part of the current Stage 1 analyses.

**Table 1.**
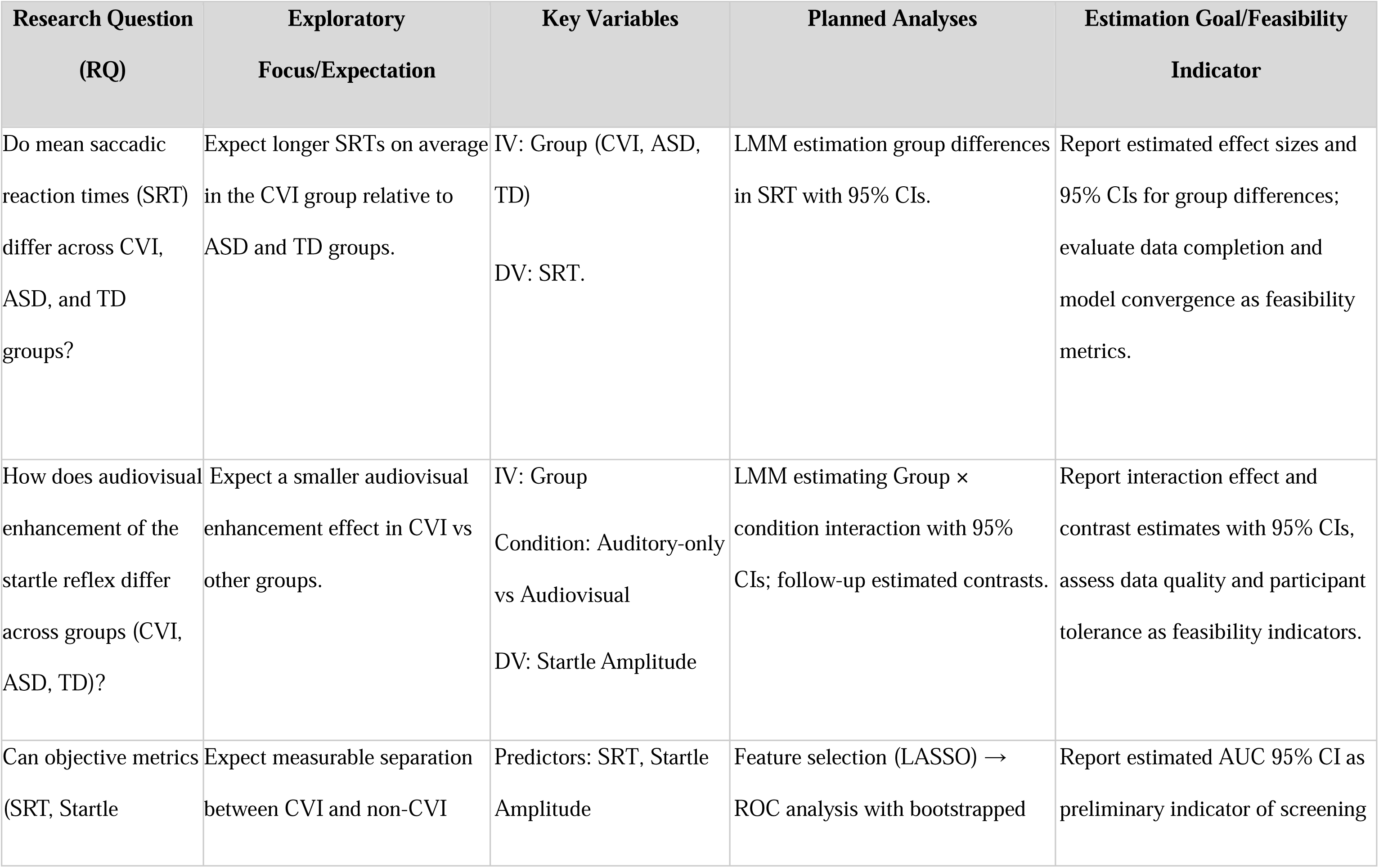
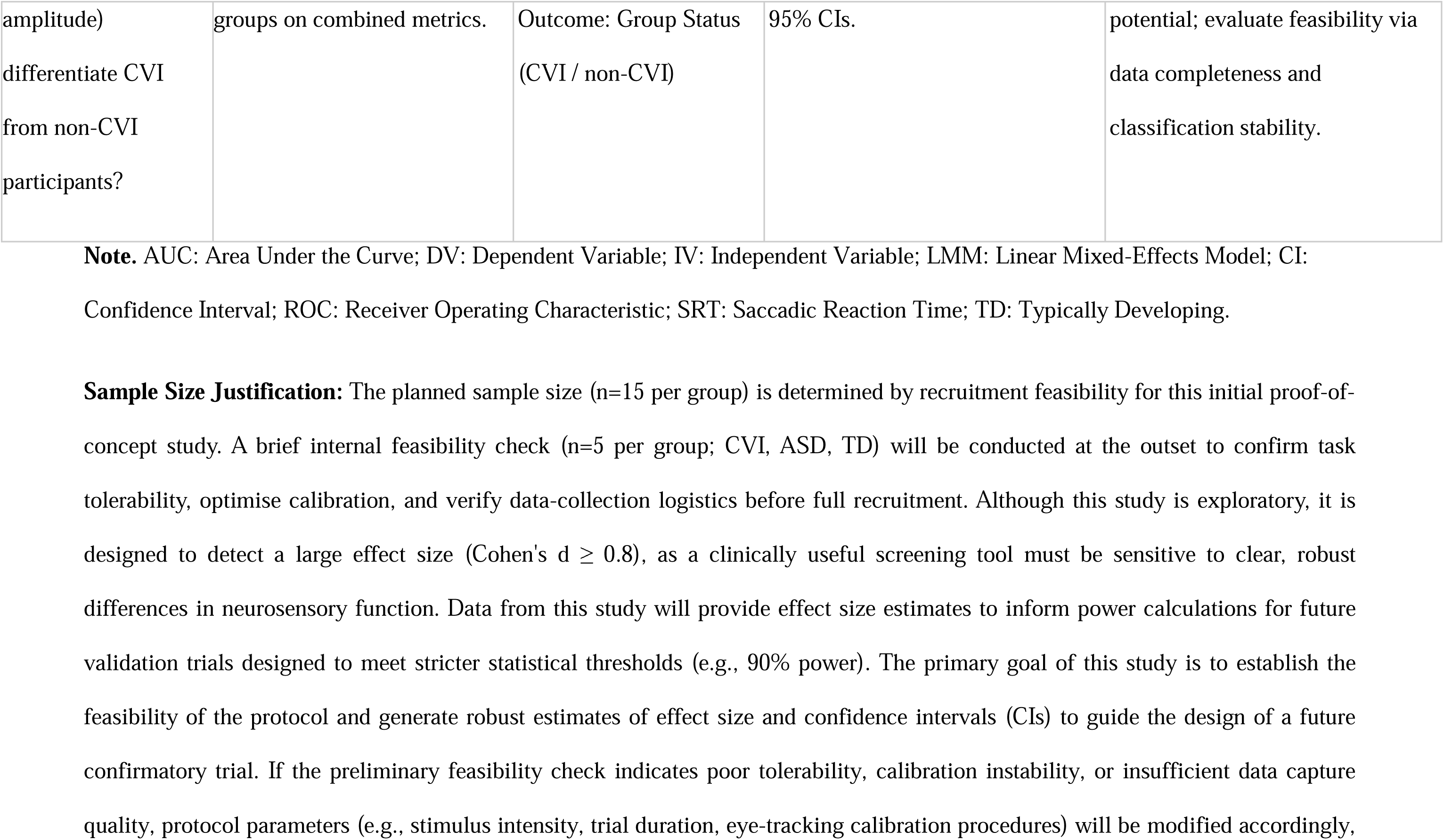
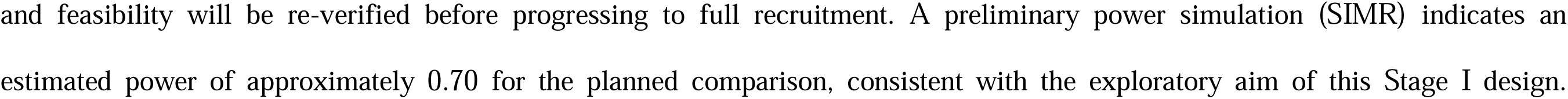
Research Questions, Variables, Planned Analyses, and Estimation Goals.

**Table 2.**
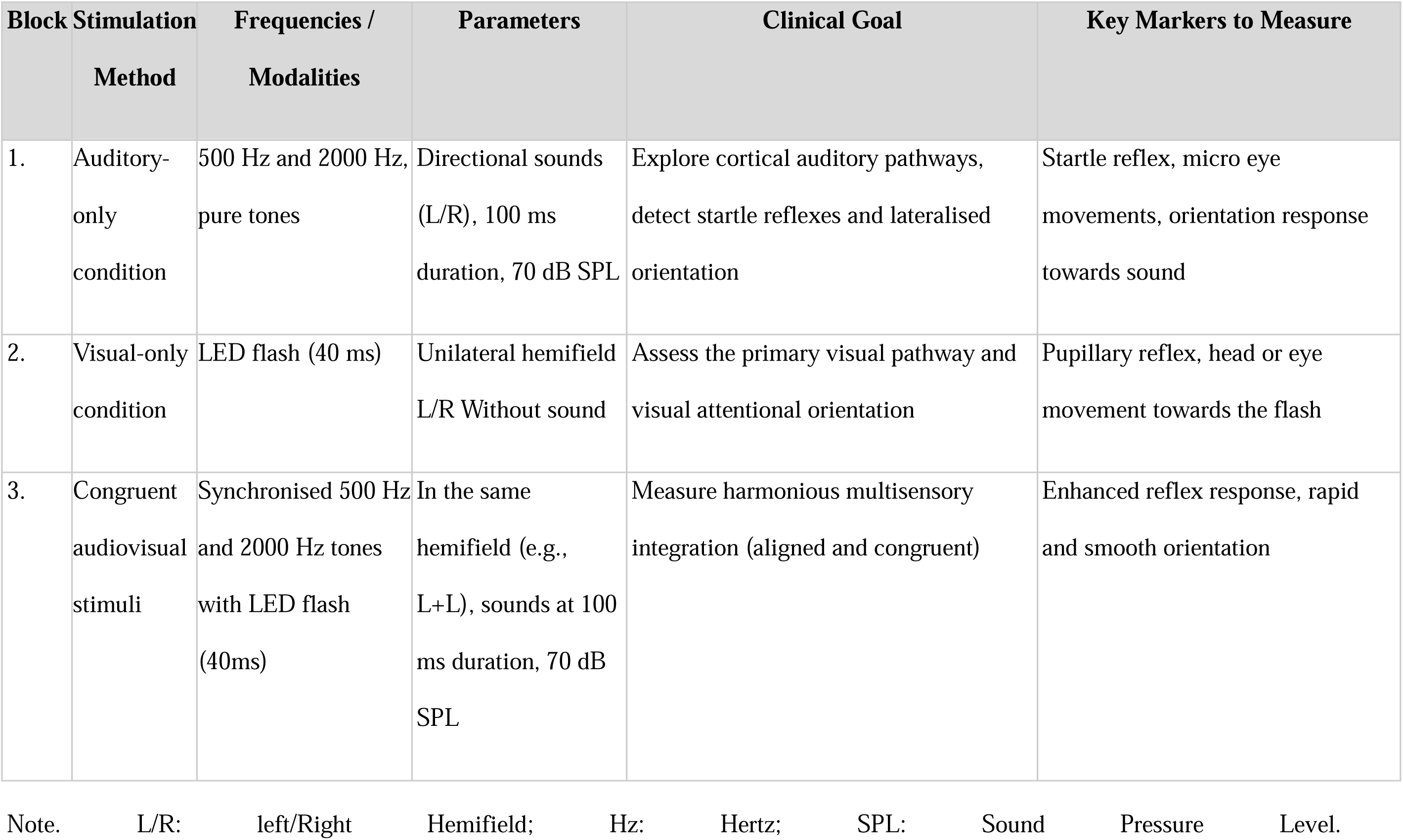
Structured Sequence of the EchoCue^TM^ Screening Protocol (Blocks for Stage 1 Analysis)

| Block | Stimulation<br>Method | Frequencies /<br>Modalities | Parameters | Clinical Goal | Key Markers to Measure |
| --- | --- | --- | --- | --- | --- |
| 1. | Auditory-only<br>condition | 500 Hz and 2000 Hz,<br>pure tones | Directional sounds<br>(L/R), 100 ms<br>duration, 70 dB SPL | Explore cortical auditory pathways,<br>detect startle reflexes and lateralised<br>orientation | Startle reflex, micro eye<br>movements, orientation response<br>towards sound |
| 2. | Visual-only<br>condition | LED flash (40 ms) | Unilateral hemifield<br>L/R Without sound | Assess the primary visual pathway and<br>visual attentional orientation | Pupillary reflex, head or eye<br>movement towards the flash |
| 3. | Congruent<br>audiovisual<br>stimuli | Synchronised 500 Hz<br>and 2000 Hz tones<br>with LED flash<br>(40ms) | In the same<br>hemifield (e.g.,<br>L+L), sounds at 100<br>ms duration, 70 dB<br>SPL | Measure harmonious multisensory<br>integration (aligned and congruent) | Enhanced reflex response, rapid<br>and smooth orientation |

288 Note. L/R: left/Right Hemifield; Hz: Hertz; SPL: Sound Pressure Level.

### Data-Based Exclusion Criteria

Pre-specified criteria will be applied to ensure data quality.

◾ **Trial-Level Exclusion:** Individual trials will be excluded from analysis if: (a) eye-tracking data quality is poor, operationally defined as loss of pupil detection for more than 50% of the trial duration, or(b) excessive head motion is detected, operationally defined as motion energy (from the mmWave radar) exceeding 3 SD from that trial’s silent baseline period. This exclusion criterion applies specifically to pre-stimulus movement. Post-stimulus motion energy peaks exceeding the same 3 SD threshold within the reflex window (40-200 ms after auditory onset) will be treated as valid startle responses rather than artefactual movement. For SRT estimation, trials will also be excluded if gaze data are missing within the critical window for detecting the first saccade (150-700 ms after target onset), rendering saccade onset unreliable. In practice, this means that trials in which gaze is lost between target onset and completion of the first saccade will be considered invalid for SRT estimation, even if total gaze loss remains below 50%. These thresholds are chosen to ensure that any analysed trial contains sufficient and valid data to reliably compute the outcome metrics.
◾ **Participant-Level Exclusion:** A participant’s entire dataset will be excluded if fewer than 60% of their total trials are deemed valid after trial-level cleaning. This threshold is set to ensure that each participant provides enough data for their individual performance to be a stable and meaningful estimate, while balancing the challenges of data collection in this population. A participant’s data will also be excluded in the event of a significant equipment malfunction (e.g., radar failure) that prevents the recording of a primary outcome for a majority of the protocol.

These thresholds are consistent with common standards in developmental eye-tracking research, where a minimum of 50% usable data per trial is often required to ensure the interpretability of gaze-based metrics (37). The 60% threshold at the participant level reflects a pragmatic trade-off between data quality and participant retention and is in line with prior studies involving young or clinical populations, where strict thresholds would otherwise lead to substantial data loss (38).

### Model Estimation and Reporting

Models will be estimated using restricted maximum likelihood (REML), with Satterthwaite approximations for degrees of freedom where required for interval estimation. Regression coefficients (ß), standard errors, and 95% CIs will be reported for all fixed effects. For interpretability, effects will be expressed both in raw units (e.g., milliseconds for SRT) and as estimated marginal means (EMMs), with 95% CIs.

Residuals will be checked for normality, homoscedasticity, and influence. Outliers beyond ±3 SD will be examined but not automatically excluded; any exclusion of outlier values will rely on a predefined, transparent rule and will be reported separately from the trial-level and participant-level exclusion criteria defined above.

Trial-level data loss due to tracking errors will be handled at the trial level, while participant-level missingness will be documented. Completion rate and usable-trial proportions will be summarised to assess data quality.

Analyses focus on estimation rather than null-hypothesis testing, with interpretation based on the direction and magnitude of effects and their 95% CIs rather than binary significance thresholds.

## Screening Accuracy Analysis

To address the third research question, whether objective metrics (SRT, Startle Amplitude) can differentiate CVI from non-CVI participants, an estimation-based approach will be used:

1. Feature selection: A principled method (e.g., LASSO regression) may be used to identify a parsimonious set of predictors (e.g., SRT, Startle Amplitude) and handle potential collinearity.
2. ROC Analysis: The performance of the final model will be assessed using Receiver Operating Characteristic (ROC) analysis. The primary outcome will be the estimated Area Under the Curve (AUC) and its 95% CIs, derived via bootstrapping with 1,000 resamples.

## Limitations and Future Directions

This study has several limitations inherent to its exploratory proof-of-concept design. First, the sample size is small and focused on a narrow age range, meaning that the results will mainly serve to generate effect size estimates for future larger-scale trials. Effect size estimates derived from small samples can be unreliable and are expected to have wide CIs; therefore, results will be interpreted with appropriate caution.

Second, the protocol relies on consumer-grade hardware, the built-in infrared camera of an iPad, for eye tracking. While this approach greatly improves accessibility, its precision for capturing subtle saccades and pupillary metrics at a typical clinical distance (e.g., 60-100 cm) in a challenging paediatric population is not yet well established in the scientific literature. Therefore, a key secondary objective of this study is to assess the feasibility of this technology. The results will provide important preliminary data on the viability of using this accessible hardware for clinical research. Future work should aim to validate these measures by comparing them to research-grade eye tracking systems.

Similarly, the use of mmWave radar to quantify startle reflex amplitude is an innovative but emerging technique for which specific validation in paediatric neurology is lacking. Therefore, a key secondary aim of this study is to provide crucial preliminary data on the feasibility and sensitivity of this method for capturing non-contact neurological reflexes in children.

Participants will be primarily recruited from specialist clinical services, which may introduce selection bias toward more severe or complex CVI cases. As a result, the sample may not fully represent the broader heterogeneity of children with CVI encountered in community or educational settings. Future studies should aim to expand recruitment to more diverse populations to improve generalisability.

The current study relies on prior CVI diagnoses provided by referring clinicians. While this reflects real-world diagnostic pathways, it introduces the risk of misclassification due to variability in diagnostic standards. This limitation is acknowledged, and future studies will include standardised diagnostic assessments and independent verification procedures to improve diagnostic reliability.

Finally, while the protocol controls for developmental level (Vineland-3), other key factors such as cognitive level (IQ) and visual acuity differences remain uncontrolled. These variables could potentially influence the results, and their absence represents a limitation of this feasibility study. Future research should aim to measure and characterise these variables more thoroughly.

## Supplementary Materials A. Optional EEG and Blocks 4-5

Note: Details regarding the optional EEG configuration and Blocks 4-5, which are not part of the present Stage 1 analyses, are provided here for completeness.

## Neural Activity Monitoring (Optional)

A flexible, front-mounted EEG headband may be integrated to record electrophysiological signals. For this proof-of-concept study, EEG use is optional to ensure that a child’s refusal to wear the headband does not become a barrier to participation. Any collected EEG data will be analysed descriptively to inform future studies.

## Experimental Blocks 4-5

These blocks include higher-level audiovisual integration tasks intended for later validation phases. They are not part of the current feasibility or estimation analyses but are documented here for transparency.

## Block 4 (Incongruent Audiovisual Stimuli)

This block introduces spatial incongruence between auditory and visual inputs (e.g., sound on the left, flash on the right) to probe the child’s ability to resolve cross-modal conflict, a process often impaired in neurodevelopmental disorders.

## Block 5 (Naturalistic Stimuli)

This block incorporates ecologically valid, semantically rich stimuli (e.g., familiar voices and animals) to assess social attention and semantic processing, thereby maintaining engagement while enabling the detection of higher-order cognitive markers.

| Block | Stimulation method | Frequencies / Modalities | Parameters | Clinical goal | Key markers to measure |
| --- | --- | --- | --- | --- | --- |
| 4. | Incongruent audiovisual stimuli | Sound and flash (1000 Hz to 3000 Hz) | Presented in opposite hemifields (e.g., L + R) | Evaluate sensory processing capacity under | Increased reaction time, hesitation or absence of |
|  |  |  |  | cross-modal<br>conflict | response |
| 5. | Naturalistic<br>stimuli | Familiar voices,<br>animal sounds<br>(250-4000 Hz)<br>synchronised<br>with animated<br>images | Familiar voices,<br>animal sounds,<br>synchronised<br>with an animated<br>image | Probe social<br>attention and<br>semantic<br>recognition | Prolonged<br>fixation,<br>smiling,<br>vocalisation,<br>motor imitation |

## Pre-Registration

This study protocol has been pre-registered on the Open Science Framework (OSF). This manuscript received Stage 1 in-principle acceptance (IPA) as a Registered Report from PCI Registered Reports.

## Declarations

### Ethics Statement

This research protocol has been developed in preparation for a future doctoral project. Formal ethics approval will be sought from the responsible Human Research Ethics Committee (HREC) at the author’s host institution prior to the commencement of any research activities involving human participants.

## Data and Materials Availability

In accordance with TOP guidelines, all processed anonymised data, analysis scripts, and digital study materials required to reproduce the study’s findings will be made publicly available on a repository such as the OSF. The EchoCue™ software script developed for stimulus delivery in this non-commercial, academic research project will be included in this public deposition.

## Data Availability Statement

The preregistration, materials, and data for this study are available on the Open Science Framework https://osf.io/yzc3b.

## Data Availability

https://osf.io/6tj3f/overview?view_only=7c5db5b7f6f24dcdaa81225ab2271e02

